# Upper Trunk Flexion Versus Whole Trunk Elevation in Semi-Fowler’s Position: Effects on Ventilation and Cardiac Chronotropy

**DOI:** 10.1101/2025.03.10.25323643

**Authors:** Sayuki Miyashita, Satoshi Kubota, Takuya Furudate

**Affiliations:** Medihos Kawaguchiko, Minamitsuru, Yamanashi, Japan; School of Health Sciences at Odawara, International University of Health and Welfare, Odarawa, Kanagawa, Japan; Graduate school, International University of Health and Welfare

**Keywords:** Semi-Fowler’s position, Tidal Volume, Minute Ventilation, RR interval, Patient Positioning, Hemodynamic Stability

## Abstract

The semi-Fowler’s position, in which the trunk is elevated 30° from the supine position, is widely used in clinical practice to manage respiratory function and reduce cardiovascular stress. Despite the known cardiovascular benefits of upper trunk flexion in the Fowler’s position, its effects in the semi-Fowler’s position remain uninvestigated. This study provides the first direct comparison of upper trunk flexion (UT30) and conventional whole-trunk elevation (WT30) at an identical 30° angle.

Fourteen healthy young women participated in a randomized crossover design comparing three positions: supine, WT30 (standard bed angle elevation), and UT30 (isolated flexion at the T10 vertebral level using cushions). Tidal volume (TV), minute ventilation (MV), respiratory rate, and RR intervals were measured after 5-minute equilibration in each position. Both UT30 and WT30 significantly increased TV (>80 mL, p<0.001) and MV (>0.9 L/min, p<0.001) compared to the supine position, with no differences between the positioning variants. Critically, RR intervals significantly decreased in the WT30 group compared to the supine position (p<0.05), indicating compensatory heart rate elevation, whereas the UT30 group maintained RR intervals equivalent to the supine position (p>0.05), demonstrating a preserved hemodynamic baseline. Although the direct comparison between UT30 and WT30 did not reach statistical significance, the distinct response patterns—hemodynamic perturbation in WT30 versus baseline stability in UT30—suggest that UT30 achieves ventilatory improvements without the associated orthostatic cardiac cost. These findings highlight UT30 as a potential strategy for decoupling respiratory support from cardiovascular stress, warranting validation in clinical populations with limited cardiac reserves.

## 1. INTRODUCTION

Fowler’s and semi-Fowler’s positions, which involve raising the trunk from the supine position, are commonly used in clinical settings to improve respiratory function and reduce cardiovascular stress (Carol A Rauen, Mary Beth Flynn Makic, 2009; Kubota et al., 2017, Kubota et al., 2015; Mary Jo Grap, 2005). Fowler’s position typically raises the trunk by 45-60°, whereas the semi-Fowler’s position involves a more moderate elevation of 30-45°. Kubota et al. demonstrated that flexing the upper trunk in the Fowler’s position resulted in less reduction in stroke volume and less increase in heart rate compared to the conventional Fowler’s position, which raises the entire trunk(Kubota et al., 2017, 2015). However, a trunk posture within the Fowler or semi-Fowler positions that minimizes respiratory stress has not been proposed.

During quiet breathing, tidal volume and minute ventilation increase when the trunk is raised(Agostoni and Hyatt, 2011a; Romei et al., 2010). This is due to the gravitational effects on the diaphragm and abdominal organs. In the supine position, gravity causes the abdominal contents to compress the diaphragm cranially, restricting diaphragmatic excursion and reducing thoracic volume. Conversely, in upright positions, gravity displaces the abdominal organs caudally, allowing greater diaphragmatic descent and increased thoracic volume. Therefore, the ventilation volume is greater in the Fowler’s position than in the supine position. Importantly, the upper trunk flexion posture proposed by Kubota et al. involves elevating the thoracic region while maintaining the abdomen in a relatively horizontal position. As the gravitational effects on the abdominal organs influence the respiratory mechanics, this differential positioning of the upper and lower trunk may produce distinct ventilatory responses. Additionally, abdominal wall tension affects respiratory function by modulating intra-abdominal pressure(Goldman et al., 1986; Novak et al., 2021). Upper trunk flexion may alter the abdominal wall tension through changes in trunk biomechanics, potentially affecting respiratory function.

Although the semi-Fowler’s position involves modest trunk elevation compared to the Fowler’s position, it is frequently employed in clinical practice. Whether upper trunk flexion within this moderate elevation range affects cardiovascular and respiratory functions remains unclear. This study aimed to examine the effects of upper trunk flexion versus conventional whole-trunk elevation in the semi-Fowler’s position on ventilation volume and RR intervals. We hypothesized that upper trunk flexion would improve respiratory function by reducing abdominal wall tension and maintaining cardiovascular stability by minimizing the hydrostatic gradients that drive orthostatic stress.

## 2. METHOD

This study employed a cross-sectional experimental design with repeated measures to compare the ventilation parameters across three different postural conditions.

### 2.1 Participants

This study was conducted in accordance with the principles of the Declaration of Helsinki and approved by the Institutional Ethics Committee of the International University of Health and Welfare (Approval No. 21-lg-16). Written informed consent was obtained from all participants before their inclusion in the study. Healthy young women were recruited using convenience sampling. The exclusion criteria were respiratory or cardiovascular diseases, low back pain, and obesity. Of the 15 participants initially measured, one was excluded due to equipment malfunction, resulting in 14 participants for analysis (age, 20.57 ± 0.73 years; height, 162.07 ± 5.4 cm; weight, 53.13 ± 5.57 kg; BMI, 20.19 ± 1.46 kg/m^2^).

This study deliberately recruited healthy young women to establish baseline physiological responses in controlled conditions. Restricting participants to a homogeneous population minimized confounding factors, such as age-related vascular compliance changes and disease-related compensatory mechanisms. Women were specifically selected given the documented sex differences in respiratory mechanics: females demonstrate a greater reliance on rib cage inspiratory muscles (Molgat-Seon et al., 2018; Sheel et al., 2016), making them an appropriate model for investigating thoracic positioning interventions.

### 2.2 Experimental Procedure

Participants were positioned in three postures: supine (SPIN) and two semi-Fowler’s positions at 30° upper trunk flexion (UT30) and 30° whole trunk elevation (WT30) (Figure 1). This study employed 30° semi-Fowler’s positions rather than Fowler’s positions exceeding 30°, as lower-angle semi-Fowler’s positions (approximately 30°) are commonly used for frail patients(Carol A Rauen, Mary Beth Flynn Makic, 2009; Grap et al., 2005; Mary Jo Grap, 2005; Zhu et al., 2020). Postures were achieved using a hospital bed, positioning cushions, and a mattress. For the UT30, trunk flexion was adjusted at the tenth thoracic vertebra (T10). The T10 level was identified by palpating the spinous processes, and cushions were placed to ensure that the flexion fulcrum was accurately located, following the method described by Kubota et al. (Kubota et al., 2015). During both the UT30 and WT30 positions, the participants were instructed to maintain their knees in slight flexion.

**Figure 1.**
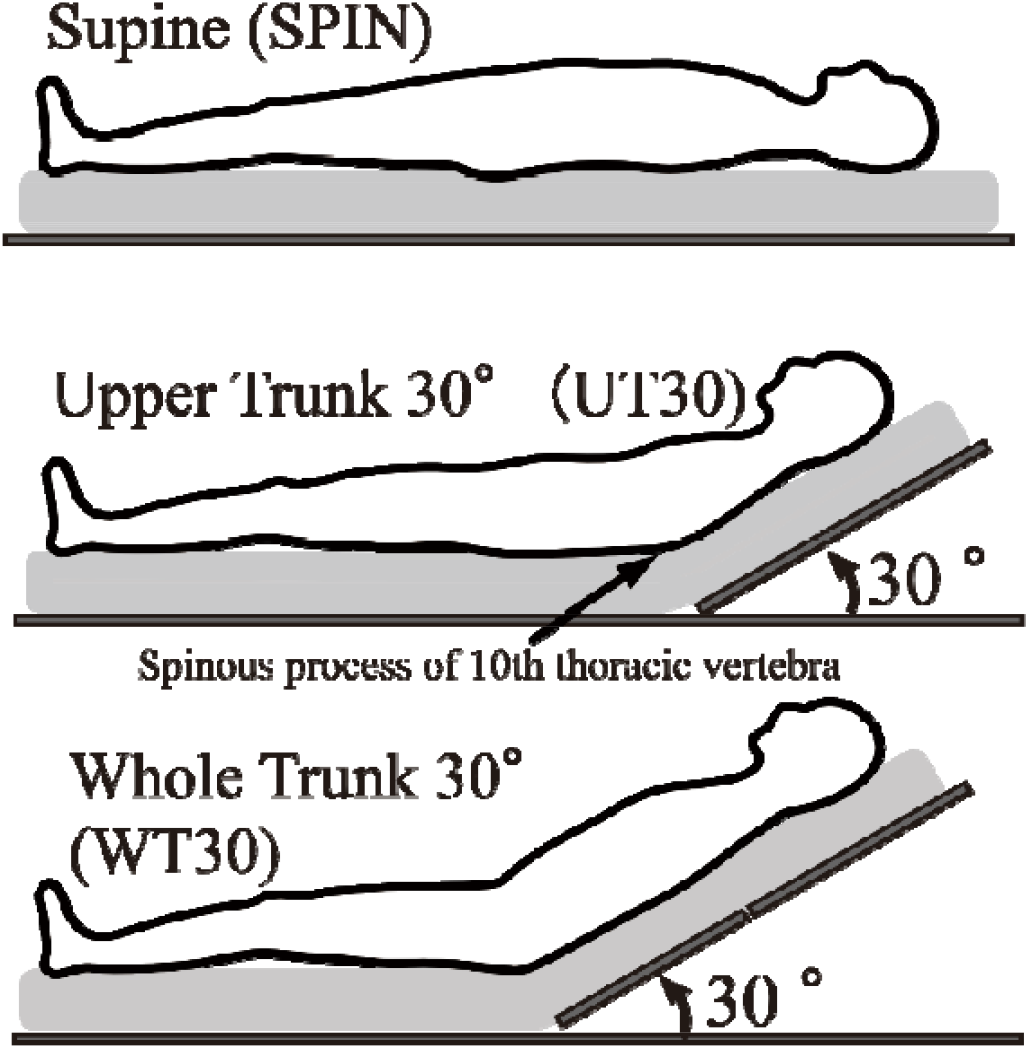
Bed positions for each condition SPIN: Supine. UT30: Lower and upper trunk inclined at 0° and 30°. Segments were subdivided based on the spinous process of 10th thoracic vertebra. WT30: Lower and upper trunk inclined at 30°.

A preliminary experiment was conducted on a separate day to allow the participants to become accustomed to breathing while wearing the ventilation measurement mask. Participants were instructed to avoid strenuous exercise, caffeine, alcohol consumption, and food intake after dinner the day before the experiment and to completely refrain from food and drink for 2 hours before the experiment. The laboratory temperature was maintained at 28°C, within the thermoneutral zone, based on previous research(Kubota et al., 2022, 2017). Participants were asked to wear sleeveless tops and shorts with properly fitted, non-constrictive bras to avoid chest compression.

All experiments were conducted between 1100 and 1500 h to minimize the circadian influences. Measurements were taken after a 15-minute rest in the supine position. Each position was measured for 5 minutes with 10-minute rest periods between measurements. The measurement order was randomized to eliminate order effects.

Ventilation volumes were measured using a ventilation measurement mask and pneumotachograph (BIOPAC Systems, TSD117, USA). Electrocardiogram (ECG) data were collected using a three-lead ECG module (BIOPAC Systems, ECG100C, USA) with electrodes placed in the standard Lead II configuration. Data acquisition was performed using a BIOPAC MP150 data acquisition system (BIOPAC Systems, CA, USA), sampled at 1000 Hz.

### 2.3 Data Analysis and Statistical Analysis

Tidal Volume (TV), Minute Ventilation (MV), Respiratory Rate (RespiR), and RR intervals (RRi) were calculated. RRi represents the time required for one cardiac cycle and corresponds to the reciprocal of the heart rate. Subjective breathing difficulty was assessed using a Numerical Rating Scale (0=easy, 10=difficult). A Linear Mixed Model (LMM) was used to analyze the effects of postures on these parameters, with postural differences as fixed effects and individual differences as random intercepts, and p-values were calculated using the Kenward–Roger method. The estimated marginal means and confidence intervals were calculated, and comparisons between postures were performed using Tukey’s method. Cohen’s d was calculated as the effect size. Statistical analyses were conducted using R version 4.3.3 (R Core Team) and the lme4 package. The significance level was set at less than 5%.

## 3 RESULTS

Table 1 presents the estimated marginal means and 95% confidence intervals for the lung volume fractions and RRi in each posture, along with the p-values and effect sizes from between-group comparisons.

**Table 1.** Respiratory Function values and RR Interval in all positions.

|  | Estimated Marginal Means(95% Confidence Intervals) |  |  | p-value(effect size) |  |  |
| --- | --- | --- | --- | --- | --- | --- |
|  | SPIN | UT30 | WT30 | SPIN vs UT30 | SPIN vs WT30 | UT30 vs WT30 |
| Tidal Volume<br>(mL) | 318.34<br>(251.26 - 385.42) | 397.06<br>(329.98 - 464.14) | 412.46<br>(345.37 - 479.54) | 0.0002<br>(1.77) | <0.0001<br>(2.11) |  |
| Minuitu Ventilation<br>(L/min) | 4.61<br>(3.69 - 5.53) | 5.5<br>(4.58 - 6.42) | 5.79<br>(4.87 - 6.71) | 0.0002<br>(1.81) | <0.0001<br>(2.39) |  |
| Respiratory Rate<br>(times/min) | 14.91<br>(13.68 - 16.13) | 14.33<br>(13.11 - 15.55) | 14.74<br>(13.52 - 15.97) |  |  |  |
| RRinterval<br>(sec) | 0.929<br>(0.884 - 0.975) | 0.908<br>(0.863 - 0.954) | 0.9<br>(0.854 - 0.945) |  | 0.0384<br>(0.99) |  |

SPIN: Supine.
UT30: Lower and upper trunk inclined at 0° and 30°.
WT30: Lower and upper trunk inclined at 30°.

TV was significantly higher in both UT30 and WT30 than in SPIN (p < 0.05), with differences exceeding approximately 80 mL in both groups. MV was also significantly higher in both UT30 and WT30 than in SPIN (p<0.05), with differences of more than 0.9 L/min. The TV and MV values were comparable between UT30 and WT30, with no significant difference. In contrast, RespiR remained consistent across all postures at approximately 14 breaths/min, and no significant differences were observed between the postures. The RRi analysis revealed distinct hemodynamic profiles between the two elevated positions. The WT30 resulted in a significantly lower RRi compared to SPIN (p<0.05), indicating a significant increase in heart rate. In contrast, UT30 showed no significant difference in RRi compared to SPIN (p>0.05), suggesting that the resting heart rate was maintained. Although the direct comparison of RRi between UT30 and WT30 did not reach statistical significance (p > 0.05), the pattern of significance relative to the baseline highlights a divergence in the cardiovascular cost. The NRS showed no significant differences between any of the postures. The effect sizes for significant between-group differences ranged from approximately 1–2.4.

## 4 DISCUSSION

This study revealed that upper trunk flexion (UT30) produced respiratory benefits comparable to whole-trunk elevation (WT30), with both positions significantly increasing TV compared to SPIN. However, the positions elicited distinct cardiovascular profiles in the participants. WT30 significantly decreased RR intervals (compensatory tachycardia), whereas UT30 maintained RR intervals similar to those in the supine condition. These findings suggest that UT30 may effectively decouple the beneficial ventilatory effects of trunk elevation from the orthostatic cardiovascular stress typically associated with it.

### 4.1 Ventilatory and Cardiovascular Functions

The significantly higher TV in WT30 than in SPIN was consistent with previous studies comparing the sitting and supine positions(Romei et al., 2010; Agostoni and Hyatt, 2011a; Takahashi et al., 1998). This can be attributed to the gravitational effect, which causes the abdominal organs to move downward when the trunk is raised, resulting in a lowered diaphragm and increased thoracic volume(Agostoni and Hyatt, 2011b; Mezidi and Guérin, 2018).

In contrast, while UT30 involves raising the thoracic region and head, the abdomen remains unraised. Therefore, the gravitational downward movement of the abdominal organs that occurs in WT30 is less likely to occur. Nevertheless, UT30 showed an increased ventilation volume compared to SPIN. This can be explained by two mechanisms. First, although the abdomen remained horizontal in UT30, upper trunk flexion altered the thoracoabdominal angle, likely reducing the cranial pressure of the abdominal organs on the diaphragm compared to the completely supine position. Second, passive support of the upper trunk likely reduces abdominal wall tension. The abdominal muscles exhibit tonic activity in upright postures but remain electrically silent in the supine position(De Troyer, 1983). Decreased abdominal wall tension reduces intra-abdominal pressure (Novak et al., 2021) and increases abdominal compliance, facilitating diaphragmatic descent (Goldman et al., 1986), even without the assistance of gravity on the viscera.

Regarding MV, which is the product of TV and RR, considering that RR remained consistent at approximately 14 breaths/min across all postures with no significant differences, we conclude that the postural differences observed in TV influenced MV, resulting in differences between SPIN and the two Semi-Fowler’s positions.

The cardiovascular responses differed markedly in their pattern relative to baseline. The significant shortening of RR intervals in WT30 reflects the expected orthostatic stress response, in which gravitational pooling reduces venous return, triggering a baroreceptor-mediated heart rate increase to maintain cardiac output. In contrast, UT30 preserved the supine RR interval values. This “hemodynamic sparing” effect is likely due to the maintenance of the lower extremities and abdomen in a horizontal plane, which minimizes the hydrostatic pressure gradient and preserves venous return. These findings are consistent with the hemodynamic results demonstrated by Kubota et al., who reported that upper trunk flexion minimized the cardiovascular burden compared with whole-trunk elevation(Kubota et al., 2017, 2015). Although the direct statistical comparison between UT30 and WT30 did not reach significance (p > 0.05), likely due to the limited sample size relative to the high autonomic reserve in healthy young participants, the qualitative difference in response patterns was physiologically critical. Specifically, WT30 induced a significant perturbation of the homeostatic baseline (significant reduction in RRi vs. SPIN), representing a distinct “physiological cost” for the gained ventilation. Conversely, UT30 did not cause such perturbations and maintained the parameters at baseline levels (hemodynamic neutrality). This divergence indicates that while WT30 imposes a cardiovascular penalty, UT30 achieves ventilatory improvement without disrupting hemodynamic stability. In clinical physiology, the maintenance of baseline versus significant perturbation represents a meaningful difference in physiological stress, suggesting that UT30 offers a superior physiological cost-benefit ratio. Although no significant differences were observed in the NRS, this experiment was conducted under resting conditions and represented a low physical burden for healthy individuals. Therefore, we believe that the participants had difficulty perceiving the differences in breathing between the different postures.

### 4.2 Clinical Implications

Upper trunk flexion in the semi-Fowler’s position represents a simple, non-pharmacological approach without specialized equipment, requiring only strategic pillow placement or adjustable bed positioning. The ability of UT30 to improve ventilation without elevating heart rate suggests a favorable “physiological cost-benefit ratio.” For healthy individuals, the increase in heart rate in WT30 is trivial. However, if these heart rate findings extend to clinical populations, patients with compromised cardiovascular function, including those with heart failure, recent myocardial infarction, or hemodynamic instability, may benefit from a positioning approach that improves ventilation without elevating the heart rate. In such vulnerable populations, the avoidance of any additional cardiovascular perturbation (as achieved in the UT30 group) is often a primary clinical goal. Supporting the clinical relevance of positioning interventions, Zhu et al. (Zhu et al., 2020) reported in a randomized clinical trial that the semi-Fowler’s position (30°) for extubation in post-abdominal surgery patients significantly reduced wound pain and severe coughing and improved patient comfort compared to the supine position. The UT30 posture, by improving ventilation without the heart rate elevation observed with WT30 in our healthy participants, might offer additional benefits to patients requiring respiratory support.

### 4.3 Limitations

This study has several limitations. First, the sample consisted of healthy young women, limiting generalizability to men, the elderly, or clinical populations with impaired autonomic reflexes. Second, while we hypothesized that reduced abdominal wall tension drives ventilatory improvement in UT30, we did not directly measure abdominal muscle activity (EMG) or intra-abdominal/gastric pressures. Future mechanistic studies should incorporate these measures to confirm the proposed pathways. Third, we measured RR intervals as a surrogate for cardiac chronotropy but did not directly assess stroke volume or cardiac output. Finally, the lack of a statistically significant difference in the direct comparison of the RRi between UT30 and WT30 warrants caution; larger studies are needed to confirm the magnitude of the hemodynamic advantage.

## 5. CONCLUSION

This study demonstrated that the semi-Fowler’s position with upper trunk flexion (UT30) improves ventilation volume comparably to whole-trunk elevation (WT30) in healthy young women. Crucially, while WT30 induced a significant elevation in heart rate (reduced RR intervals) relative to the supine position, UT30 maintained the heart rate at baseline levels. This divergence indicates that while both positions offer respiratory benefits, they carry different physiological costs; WT30 imposes a hemodynamic penalty that UT30 avoids. These findings suggest that UT30 provides a distinct physiological advantage by uncoupling the ventilatory support from orthostatic cardiovascular stress. This posture warrants further investigation as a strategic alternative for hemodynamically fragile patients requiring respiratory support.

## Acknowledgment

We would like to thank the participants of our study and Professor Sumiko Yamamoto and Professor Shinichiro Ishii for their valuable assistance.

## Author Contributions

Conceptualization: Sayuki Miyashita, Satoshi Kubota; Data curation: Sayuki Miyashita, Satoshi Kubota, Takuya Furudate; Formal analysis: Satoshi Kubota, Takuya Furudate; Investigation: Sayuki Miyashita, Satoshi Kubota, Takuya Furudate; Methodology: Satoshi Kubota, Sayuki Miyashita; Project administration: Sayuki Miyashita, Satoshi Kubota; Supervision: Satoshi Kubota; Writing—original draft: Satoshi Kubota; Writing—review & editing: Sayuki Miyashita, Takuya Furudate.

## Data Availability Statement

The authors will make the data supporting the conclusions of this article available upon reasonable request.

## References

Agostoni, E., Hyatt, R.E., 2011a. Static Behavior of the Respiratory System. Compr. Physiol., Major Reference Works. doi:10.1002/cphy.cp030309

Agostoni, E., Hyatt, R.E., 2011b. Static Behavior of the Respiratory System, in: Comprehensive Physiology. pp. 113–130. 10.1002/cphy.cp030309

Carol A Rauen, Mary Beth Flynn Makic, E.B., 2009. Evidence-based practice habits: transforming research into bedside practice. Crit Care Nurse 29(2), 46–59.

De Troyer, A., 1983. Mechanical role of the abdominal muscles in relation to posture. Respir. Physiol. 53, 341–353. 10.1016/0034-5687(83)90124-X

Goldman, J.M., Silver, J.R., Lehr, R.P., 1986. An electromyographic study of the abdominal muscles of tetraplegic patients. Spinal Cord 24, 241–246. 10.1038/sc.1986.33

Grap, M.J., Munro, C.L., Hummel, R.S., III, Elswick, R.K., Jr, McKinney, J.L., Sessler, C.N., 2005. Effect of Backrest Elevation on the Development of Ventilator-Associated Pneumonia. Am. J. Crit. Care 14, 325–332. 10.4037/ajcc2005.14.4.325

Kubota, S., Endo, Y., Kubota, M., Ishizuka, Y., Furudate, T., 2015. Effects of trunk posture in Fowler’s position on hemodynamics. Auton. Neurosci. Basic Clin. 189, 56–59. 10.1016/j.autneu.2015.01.002

Kubota, S., Endo, Y., Kubota, M., Miyazaki, H., Shigemasa, T., 2022. The Pressor Response to the Drinking of Cold Water and Cold Carbonated Water in Healthy Younger and Older Adults. Front. Neurol. 12.

Kubota, S., Endo, Y., Kubota, M., Shigemasa, T., 2017. Assessment of effects of differences in trunk posture during Fowler’s position on hemodynamics and cardiovascular regulation in older and younger subjects. Clin. Interv. Aging 12, 603–610. 10.2147/CIA.S132399

Mary Jo Grap, C.L.M., 2005. Quality improvement in backrest elevation: improving outcomes in critical care. AACN Clin Issues 16(2), 133–9.

Mezidi, M., Guérin, C., 2018. Effects of patient positioning on respiratory mechanics in mechanically ventilated ICU patients. Ann. Transl. Med. 6, 1–9.

Molgat-Seon, Y., Peters, C.M., Sheel, A.W., 2018. Sex-differences in the human respiratory system and their impact on resting pulmonary function and the integrative response to exercise. Sex Differ. 6, 21–27. 10.1016/j.cophys.2018.03.007

Novak, J., Jacisko, J., Busch, A., Cerny, P., Stribrny, M., Kovari, M., Podskalska, P., Kolar, P., Kobesova, A., 2021. Intra-abdominal pressure correlates with abdominal wall tension during clinical evaluation tests. Clin. Biomech. 88, 105426. 10.1016/j.clinbiomech.2021.105426

Romei, M., Mauro, A. Lo, D’Angelo, M.G., Turconi, A.C., Bresolin, N., Pedotti, A., Aliverti, A., 2010. Effects of gender and posture on thoraco-abdominal kinematics during quiet breathing in healthy adults. Respir. Physiol. Neurobiol. 172, 184–191. 10.1016/j.resp.2010.05.018

Sheel, A.W., Dominelli, P.B., Molgat-Seon, Y., 2016. Revisiting dysanapsis: sex-based differences in airways and the mechanics of breathing during exercise. Exp. Physiol. 101, 213–218. 10.1113/EP085366

Takahashi, T., Yamada, S., Tanabe, K., Nakayama, M., Osada, N., Itoh, H., Murayama, M., 1998. The Effects of Posture on the Ventilatory Responses During Exercise. J. Jpn. Phys. Ther. Assoc. 1, 13–17. 10.1298/jjpta.1.13

Zhu, Q., Huang, Z., Ma, Q., Wu, Z., Kang, Y., Zhang, M., Gan, T., Wang, M., Huang, F., 2020. Supine versus semi-Fowler’s positions for tracheal extubation in abdominal surgery-a randomized clinical trial. BMC Anesthesiol. 20, 185. 10.1186/s12871-020-01108-5

